# An Initial Psychometric Evaluation of the APS-POQ-R in Acute Pain Presenting to the Emergency Department

**DOI:** 10.1101/2020.09.15.20194738

**Authors:** James A Hughes, Lee Jones, Joseph Potter, Alixandra Wong, Nathan J Brown, Kevin Chu

**Affiliations:** Emergency and Trauma Centre, Royal Brisbane and Women’s Hospital, Brisbane, Australia; School of Nursing, Queensland University of Technology, Brisbane, Australia; Institute of Health and Biomedical Innovation, Queensland University of Technology, Brisbane, Australia; Logan Hospital, Meadowbrook, Australia.; Faculty of Medicine, University of Queensland, Brisbane, Australia.

## Abstract

3.

**Background:** Pain is a common presenting complaint to the emergency department (ED), yet is often undertreated. When assessing the outcomes of pain care in the ED, process measures are commonly reported. Attempts to measure patient-reported outcomes existing in current ED literature. However, they are frequently unvalidated and lack standardization. The American Pain Societies – Patient Outcome Questionnaire-Revised edition (APS-POQ-R) has been identified as the most likely, pre-existing tool to be useful in the acute pain in the ED. However, this requires feasibility and construct validation before use.

**Objective:** To assess the feasibility and construct validity of the APS-POQ-R in patients presenting to the adult emergency department with acute pain.

**Methods:** This study is an initial psychometric evaluation of the constructs contained within the APS-POQ-R in adult patients presenting with moderate to severe acute pain to a large urban ED. The study is guided by the methods described in the initial development of the instrument.

**Results:** Two hundred adult patients were recruited and completed the APS-POQ-R. The APS-POQ-R demonstrated content validity in patients presenting with acute pain. Exploratory factor analysis demonstrated five subgroups. The tool demonstrated discriminatory ability based on patient urgency, and subscale measurement was associated with patient satisfaction with care.

**Conclusions:** The APS-POQ-R has demonstrable construct validity in adult patients presenting with acute pain to the ED. Further psychometric analysis across multiple EDs is required before the APS-POQ-R can be recommended as a validated PROM for ED patients in pain.

## 4. Introduction

Up to 70% of all patients presenting to the emergency department (ED) will have pain^1,2^. Undertreatment of pain in ED patients has long been recognized^3^ and is highly likely to occur in specific patient subgroups, such as those with cognitive impairment^4^. On the other hand, EDs are frequently criticized for overtreating patients with opiates^5^ and strategies to reduce the amount of opiate prescribing within EDs and on discharge are common^6^. While the precise reasons for undertreatment and overtreatment of pain in EDs are unknown, one possible reason is the lack of suitable and validated patient-reported outcome measures (PROMs) to guide care.

Pain is a subjective experience. The outcomes of pain care are best measured from the perspective of the patient^7^. Ideally, the outcome should not require interpretation by the clinician as significant bias can occur when clinicians evaluate pain severity and response to treatment^8^. Other factors, such as gender may influence factors such as stress and anxiety associated with pain, that in turn influence the outcomes of pain care^9^. Alterations in pain intensity do not correlate well with patient-reported analgesia^10^.

Furthermore, statistically significant changes in pain intensity may not correlate to clinically meaningful patient-reported changes as such scores are not sensitive to small changes^11^. For these reasons, many practitioners who care for patients in pain, or at risk of pain, rely on PROMs that take into account the multi-faceted nature of pain and analgesia. In ED pain research, PROMs are uncommon and, where they have been used, are most likely to be measures of satisfaction with care in the ED setting^12^.

In the absence of validated PROMs for pain care in the ED setting, clinicians and researchers have had to rely on the system- or process-based outcome measures to help evaluate and compare treatments. One example of these is the time it takes until the first analgesic medication is administered. However, the usefulness of such measures in assessing quality or effectiveness of pain care is presumed, in this case, that “faster is better”, that may not necessarily hold^13,14^. Another commonly used outcome measure is a pain intensity rating. A drop of two points on an eleven-point scale until the patient rates their pain less than a four is considered to be clinically significant and represent achieving “adequate analgesia.”^15,16^. While pain intensity ratings can be regarded as patient-related, they are somewhat unidimensional and fail to capture the subjective experiences associated with pain and analgesia. The uniqueness of patient-reported outcome measurement in research is that measurements come directly from the patient, without interpretation of the clinician or researcher^17^. In the absence of objective measures of pain, and with only a small amount of correlation between time based metrics and pain treatment^18^ patient-reported outcomes and multi dimentional PROMs are useful in capturing the patients experience of illness and treatment, health systems performance and may have prognostic signifigance^17^.

Validating existing pain care PROMs for use in the ED setting would give researchers the tools with which to compare treatments and improve ED pain care. While several patient-reported outcomes have been used in ED pain research^12^, only one has been explicitly validated for use in the ED: a Danish translation of the American Pain Society– Patient Outcome Questionaire-Revised Edition (APS-POQ-R)^19^. The validity of the tool, however, is limited by its validation in patients with acute abdominal pain only^19^. Despite this, the APS-POQ-R is the most promising PROM for measuring patient-reported outcomes of pain care in the adult ED, and comprehensive psychometric testing is required to establish the validity of an English version in the broad spectrum of ED patients with acute pain^12^.

The objective of the current study was to examine the psychometric properties of the English version of APS-POQ-R in adult patients presenting with moderate to severe acute pain to an ED. The specific aims were: 1) to determine the feasibility of administering the APS-POQ-R to adult patients after completion of their care, before leaving the ED, and 2) to determine the construct validity of the APS-POQ-R in measuring patient-centred outcomes of acute pain care.

## 5. Methods

This prospective psychometric evaluation of the APS-POQ-R in adult ED patients with moderate to severe acute pain was conducted at the Emergency and Trauma Centre at Royal Brisbane and Women’s Hospital, Australia and approved by the hospital’s Human Research Ethics Committee (LNR/2019/QRBW/55143). All participants provided written informed consent.

### The Instrument

We used a modified version of the APS-POQ-R. The APS-POQ-R is designed to evaluate care within a hospital-based quality improvement or research framework. It measures six aspects of quality care including 1) pain severity and relief,

2) impact of pain, 3) side effects of treatment, 4) information regarding pain relief and treatment, 5) ability to participate in decision making about pain care and 6) the use of non-pharmacological strategies^20^. Our modifications of the APS-POQ-R were necessary to make it more suitable for ED use. These modifications were minor and related to the timeframe the patient was asked to report on and the location. Specifically, the APS-POQ-R questions that had previously used the phrase *“The following questions are about pain you experienced during the first 24 hours in the hospital or after your operation”* were modified to *“The following questions are about pain you experienced in the emergency department”*. Questions that used the phrase *“in the first 24 hours”* were modified to “*in the emergency department”*. (See supplemental material for survey design).

### Sample

All adult patients with moderate to severe acute pain were eligible to participate. Eligible patients were identified through the ED Information System (EDIS). Patients were invited to participate after their care was completed and before they left the ED if they met the following screening criteria:

- They had presented with pain greater than 3/10 on arrival;
- They were experiencing pain that has been present for less than six weeks.
- They were able to understand and communicate in English;
- They were cognitively intact and able to provide written informed consent.

Care was deemed to be complete when the patient was ready for discharge home or had a discharge destination identified in EDIS, such as admission to ED short stay or an inpatient unit. Patients were also recruited from the ED short stay unit, but they were not approached after they had already left the ED. Patients who had been transferred to the ED from another hospital were ineligible for participation. Patients on repeat ED visits, who had previously participated in the study, were not recruited a second time. There are 18 questions in the APS-POQ-R, therefore 200 patients were recruited to meet the ten observations per variable criteria and allow for up to 5% missing data^21^.

### Data Collection

Data were collected over eight, non-consecutive weeks between August and December 2019 by two medical students who were part of the investigator team. Consenting patients were interviewed by one of the medical students, and the patient’s answers were entered directly into a REDCap (Vanderbilt University) database. Additional data about the presentation and treatment given in the ED were collected from the patient’s electronic medical record.

### Analysis

The statistical analysis and psychometric evaluation are consistent with the work of Gordon et al. (2010) in which the APS-POQ-R was developed. This methodology has been used in the psychometric evaluation of the APS-POQ-R in other populations as described by Botti et al., Schultz et al., Zoega et al., and Rothuag et al. ^20,22^–^25^.

### Descriptive Statistics

Patient and treatment variables are reported using means and standard deviations, and categorical variables are summarized using frequencies and percentages. Results of the items on the APS-POQ-R are presented as minimum, maximums and means of all questions, including non-response rates.

### Missing data

For participants with item-response rates of less than 70%, the records were deemed incomplete and omitted from further analysis. For participants with item-response rates up to 30%, the non-responses were imputed using expectation maximization^26^ and the records, including imputed values, were included in the analysis.

### Internal Consistency

The internal consistency of the tool as a whole and for each subscale were calculated and presented using Cronbach’s alpha. Subscale item to total correlations is also presented as a Cronbach’s alpha if the item was deleted from the scale. Subscale means and variance are presented for each of the items in each scale and if they were deleted. Item-to-item correlations within each subscale that are moderately or highly correlated (correlation higher the 0.4) were also flagged.

### Construct Validity

Construct validity to the degree to which a test measures what it claims, or purports to measure^27^. Exploratory factor analysis (principal axis factoring) was used to explore the items corresponding to latent factors from the tool. Previously Gordon et al. described five factors within the tool. The number of factors chosen is based on the proportion of variance explained by eigenvalues and coherency of the underlying factors to form clinical constructs, and this is represented graphically on the Scree plot. All questions loaded to at least one factor with a weight of 0.3 or higher; therefore, all questions were retained in the analysis. A Kaiser-Meyer-Olkin (KMO) measure of sampling adequacy higher than 0.6 was considered adequate^28,29^. A Bartlett’s test of sphericity was used to detect at least one significant correlation, and this was considered significant if the p-value was less than 0.05^30^.

### Contrasting Groups

There have been several contrasting groups previously described in ED pain care literature. Gender^31^, age^32^, socio-economic status^18^, triage score^33^ and the timeliness of the administration of analgesic medication^13^ all influence ED pain care. To assess these contrasting groups in terms of patient satisfaction and patient participation in decisions about their treatment, differences between these groups were assessed using t-tests.

## 6. Results

A total of 264 patients were identified as eligible for the study, of which 200 (76%) consented into the study. Of the 64 patients not recruited: 35 refused consent, 13 were discharged before the invitation to participate, one patient withdrew consent during the interview, 12 patients reported pain less than or equal to 3/10, and two patients were physically unable to consent.

The characteristics of the participants are summarised in Table One. The mean age of the participants was 43 (95% confidence interval (CI): 40.5–45.5) years. Participants were distributed throughout all urgency categories (Australasian Triage Score (ATS) categories 1 to 5), with most participants in the ATS 3 (to be seen within 30 minutes) and 4 (to be seen within one hour) categories. Participants had a mean Index of Relative Socio-economic Advantage and Disadvantage (IRSAD) score of 1057, which indicates slightly higher affluence compared with the average Australian population (IRSAD of 1000)^34^. Private health insurance was held by 42.5% of participants. Participants had a low burden of comorbidities, as reflected in the median Charlson score of ^35^. Participants arrived with pain of all aetiologies, except for cardiac chest pain. The most common forms of pain were abdominal/genitourinary pain 83 (41.5%), fractures 35 (17.5%), musculoskeletal injuries 29 (14.5%). Participants arrived with a median pain score of 8.0 (Interquartile range 7.0 – 9.0) out of 10 on a numerical pain rating scale.

**Table One:** Description of the participants.

| Variable |  | Result |
| --- | --- | --- |
| Age (mean, SD) |  | 43 years (17.9 years) |
| Sex (n, %) | Male | 91 (45.5%) |
|  | Female | 109 (54.5%) |
| ATS (n, %) | One (Immediate) | 1 (0.5%) |
|  | Two (within 10 minutes) | 27 (13.5%) |
|  | Three (within 30 minutes) | 89 (44.5%) |
|  | Four (within 60 minutes) | 76 (38.0%) |
|  | Five (within 120 minutes) | 7 (3.5%) |
| IRSAD (mean, SD) |  | 1057 (63.2) |
| Private Health Insurance (n, %) | Yes | 85 (42.5%) |
|  | No | 115 (57.5%) |
| Charlson Score (median, IQR) |  | 0 (0 – 2) |
| Country of Birth (n, %) | Australia | 144 (72.0%) |
|  | Not Australia | 54 (27.0%) |
| Mode of Arrival (n, %) | Ambulance Service | 88 (44.0%) |
|  | Walk-in | 112 (56.0%) |

SD = Standard Deviation; ATS = Australasian Triage Score, with maximum waiting times for medical assessment and treatment; IRSAD = Index of Relative Socioeconomic Advantage and Disadvantage (>1000 = advantaged ; <1000 disadvantaged). Charlson Score = higher the number the greater the burden of comorbid disease and the greater the change of death over the next ten years.

### Means, Standard Deviations, and Frequencies of the APS-POQ-R questions

The APS-POQ-R, as described by Gordon et al., had 18 items measured on a 10-point continuous scale. Due to a transcription error to the electronic database, only 17 of these items were asked of the 200 participants in this study, with the omitted item being related to how the participant’s pain caused them to feel anxious. The following table (Table Two) describes the minimum, maximum, mean and standard deviation for each of the 17 items.

**Table Two:** Description of the responses from the first nine questions of the APS-POQ-R.

|  | Question | N | Minimum Response | Maximum Response | Mean Response | SD |
| --- | --- | --- | --- | --- | --- | --- |
| 1. | On this scale, please indicate the least pain that you had in the emergency department. | 200 | 0 | 10 | 4.99 | 2.97 |
| 2. | On this scale, please indicate the worst pain you had in the emergency department. | 200 | 4 | 10 | 7.94 | 1.61 |
| 3. | How often were you in severe pain in the emergency department? Please select the best estimate of the percentage of time you experienced severe pain | 200 | 10% | 100% | 63.2% | 36.6% |
| 4. | Select one number that best describes how much pain interfered or prevented you from: |  |  |  |  |  |
|  | a) Doing activities in bed such as turning, sitting up, repositioning? | 200 | 0 | 10 | 6.73 | 3.49 |
|  | b) Doing activities out of bed, such as walking, sitting in a chair, standing at a sink? | 200 | 0 | 10 | 6.58 | 3.74 |
|  | c) Falling asleep? | 199* | 0 | 10 | 6.85 | 3.91 |
|  | d) Staying asleep? | 198* | 0 | 10 | 6.80 | 3.99 |
| 5. | Pain can affect our mood and emotions. On this scale, please select the one number that best shows how much the pain caused you to feel: |  |  |  |  |  |
|  | a) Depressed? | 198* | 0 | 10 | 2.94 | 3.73 |
|  | b) Frightened? | 198* | 0 | 10 | 2.60 | 3.66 |
|  | c) Helpless? | 197* | 0 | 10 | 3.97 | 3.95 |
| 6. | Have you had any of the following side effects? Please select 0 if no. Please circle the number that best shows the severity of each: |  |  |  |  |  |
|  | a) Nausea? | 197* | 0 | 10 | 2.23 | 3.51 |
|  | b) Drowsiness? | 197* | 0 | 10 | 2.37 | 3.45 |
|  | c) Itching? | 195* | 0 | 10 | 0.38 | 1.69 |
|  | d) Dizziness? | 197* | 0 | 10 | 1.55 | 2.88 |
| 7. | In the emergency department, how much relief of your pain did you receive? Please circle the one percentage that best shows how much relief you have received from all of your pain treatments combined? | 193* | 0 | 10 | 5.75 | 3.05 |
| 8. | Were you allowed to participate in decisions about your pain treatment as much as you wanted to? | 198* | 0 | 10 | 9.24 | 2.24 |
| 9. | Select one number that best shows how satisfied you are with the results of your pain treatment in the emergency department? | 195* | 0 | 10 | 8.91 | 1.98 |

\* Numbers less than 200 indicate missing responses to these questions. 16 (8%) of respondents had missing answers. SD = Standard Deviation. All items were measured on a scale of 0 (0%) to 10 (100%) where 0 equalled a positive outcome (i.e. no pain) to 10 equating to a negative outcome (i.e. worst possible pain).

### Missing data

Only 16 (8%) participants had non-response items in their surveys. The questions with missing responses are outlined in Table Two. Missing responses were assessed by Little’s test to assess if they met the missing completely at random (MCAR) assumption. Little’s test indicated that the missing vales did not violate the MCAR assumption (ᵡ^2^ = 134.4 (143), p=0.685). Four participants missed answers to more than 30% of the questions and were excluded from further analysis. The remaining 12 participants with missing responses had their missing data imputed through expectation maximization^36^.

### Exploratory Factor Analysis

Exploratory factor analysis (principal axis factoring) with Promax rotation and Kaiser Normalisazion was applied to the 17 questions asked that had been previously described. Six identifiable factors had Eigenvalues greater than one (range: 3.47 – 1.02), accounting for 65.8% of the variation seen in the responses. All questions mapped to a factor with a factor loading of at least 0.3. There was very little difference between a five-factor solution and a six-factor solution with only 6% of additional variation explained by a six-factor solution. The six-factor solution had several issues, including increasing the number of questions that mapped to factors below 0.4 but higher than 0.3, two factors that had only two questions, and splitting of the pain severity and interference factor. As can be seen by the scree plot in Figure One the point of inflection of the Eigenvalues greater then one is at five factors, not six. Therefore, in line with previous APS-POQ-R, validation studies, a five subscale solution is reported.

**Figure One:**
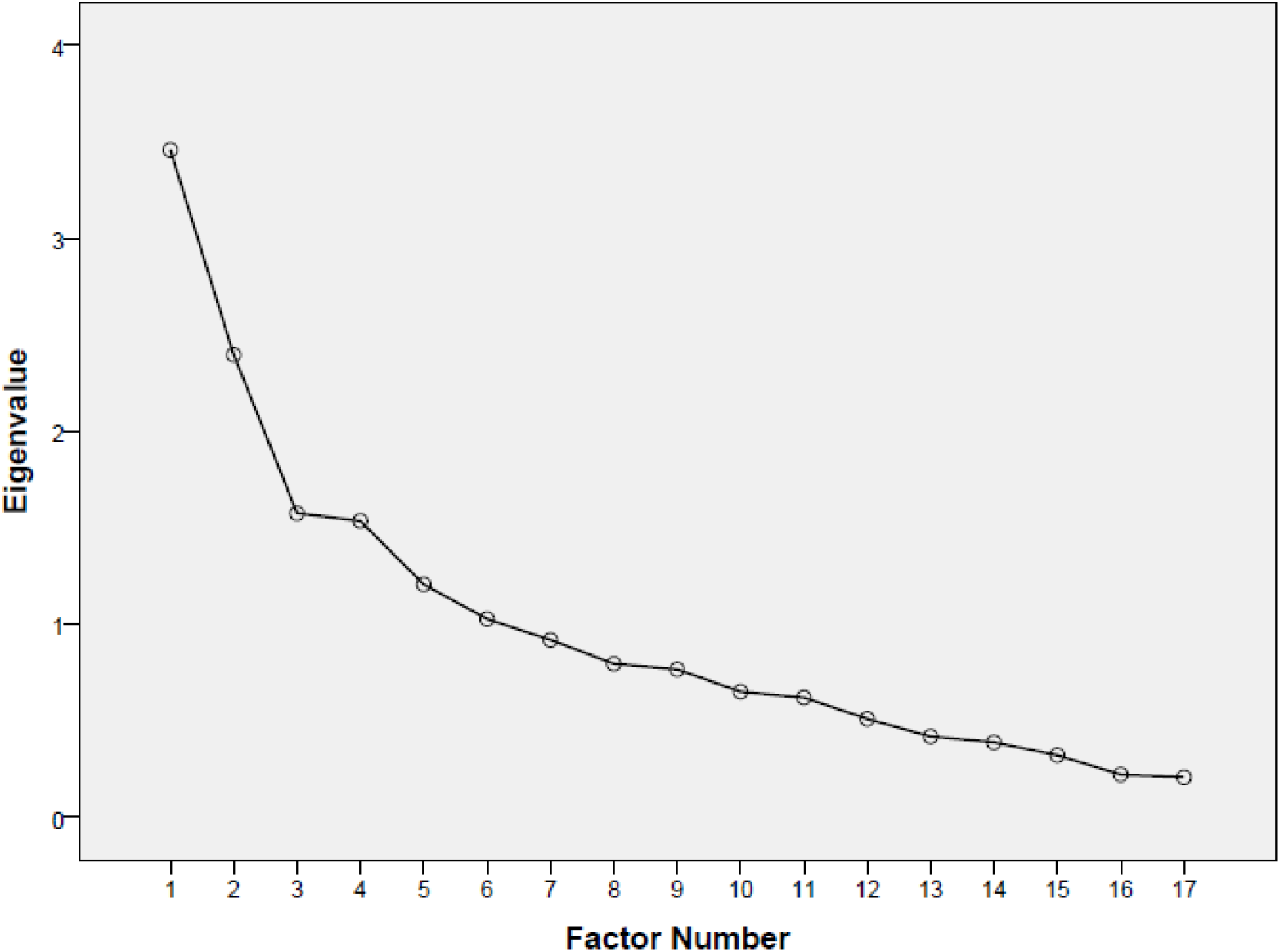
Scree plot of the Eigenvalues for each number of factors from one to seventeen

Table Three shows the factor loading for a five subscale solution that explains 59.85% of the variation in the measure. One item, participation in the decision about your pain treatment, loads to a factor below 0.4. However, inclusion of the patient in a patient-centred approach to pain outcome measurement was one of the main objectives of validating this tool; therefore, it was retained in the instrument. Factor one represents pain severity and activity interference subscale, factor two represents the affective subscale, factor three is the sleep interference subscale, factor four the adverse effects subscale and factor five is the perceptions of care subscale.

**Table Three:** Pattern Matrix

|  |  | Factor |  |  |  |  |
| --- | --- | --- | --- | --- | --- | --- |
|  |  | 1 | 2 | 3 | 4 | 5 |
| 1. | On this scale please indicate the least pain that you had in the emergency department? | .676 | -.163 | .163 | -.010 | .114 |
| 2. | On this scale please indicate the worst pain you had in the emergency department? | .409 | .289 | .063 | .069 | -.128 |
| 3. | How often were you in severe pain in the emergency department? | .555 | -.340 | .072 | -.057 | .184 |
| 4. | Select one number below that best describes how much pain interfered or prevented you from: | .610 | .171 | .032 | -.020 | -.085 |
|  | A. Doing activities in bed such as turning, sitting up, repositioning |  |  |  |  |  |
|  | B. Doing activities out of bed such as walking, sitting in a chair, standing at sink | .847 | .185 | -.182 | .010 | -.139 |
|  | C. Falling asleep | .066 | .106 | .739 | -.007 | -.045 |
|  | D. Staying asleep | -.042 | .066 | .980 | -.007 | -.014 |
| 5. | Pain can affect our mood and emotions. On this scale, please select the one number that best shows how much the pain caused you to feel: |  |  |  |  |  |
|  | A. Depressed | -.114 | .595 | .169 | .084 | .080 |
|  | B. Frightened | .026 | .579 | .036 | -.111 | .103 |
|  | C. Helpless | .330 | .717 | -.035 | -.006 | .122 |
| 6. | Have you had any of the following side effects? Please select "0" if no; if yes, please circle the number that best shows the severity of each. |  |  |  |  |  |
|  | A. Nausea | .238 | -.144 | .064 | .433 | -.019 |
|  | B. Drowsiness | -.144 | .128 | .007 | .544 | .092 |
|  | C. Itching | -.075 | .132 | -.016 | .421 | -.187 |
|  | D. Dizziness | .106 | -.159 | -.043 | .652 | .108 |
| 7. | In the emergency department, how much relief of your pain did you receive? | -.062 | .106 | .014 | -.006 | .430 |
| 8. | Were you allowed to participate in decisions about your pain treatment as much as you wanted to? | .004 | .007 | -.010 | .038 | .365 |
| 9. | Select one number that best shows how satisfied you are with the results of your pain treatment in the emergency department. | -.083 | .258 | -.086 | .022 | .723 |

Factor 1: Pain Severity and Interference Subscale; Factor 2: Sleep Subscale; Factor 3: Affective Subscale; Factor 4: Side-Effects subscale; Factor 5: Perceptions of Care subscale.

Table Four shows the correlation between the subscales. While none of the subscales is highly correlated with each other, predictably the pain severity and interference subscale are moderately correlated with the affective subscale. This demonstrates that each subscale is measuring a distinct aspect of the patient-reported outcomes of pain care.

**Table Four:** Factor correlation matrix

|  | **Pain Severity and Interference subscale** | **Sleep subscale** | **Affective subscale** | **Side-Effects subscale** | **Perceptions of Care Subscale** |
| --- | --- | --- | --- | --- | --- |
| **Pain Severity and Interference Subscale** | 1.000 |  |  |  |  |
| **Sleep subscale** | -.056 | 1.000 |  |  |  |
| **Affective subscale** | .469 | .004 | 1.000 |  |  |
| **Side-Effects subscale** | .171 | .282 | .101 | 1.000 |  |
| **Perceptions of Care subscale** | .100 | .025 | -.021 | .030 | 1.000 |

The affective, adverse effects and perceptions of care subscales map to the same items that have previously been reported by Gordon et al., (2010). This evaluation has shown that pain severity and the completion of activities in and out of bed are more closely related, and map to the same subscale. In the previous evaluation of this instrument pain severity and sleep have mapped to the same subscale. In the ED population sleep maps to its own subscale (see Table Five).

### Item Correlations and Reliability

The subscale means, variance, item to scale correlations, and internal consistency of each of the subscales is presented in Table Five. This table includes subscale totals and internal consistency if each item were deleted from each subscale. Pain severity and interference, sleep and affective subscales have adequate levels of internal consistency with side effects and perception of care having lower levels of internal consistency. Item-to-item correlations were assessed within each subscale, with any item-to-item that demonstrated a moderate correlation (greater than rho = 0.4) being considered significant^37^. Within the pain severity and interference subscale, there was a high inverse correlation between the time the participant reported being in severe pain and the least pain they reported while in the ED (r = −0.684, p< 0.001). There was an equally high correlation between the pain interference for activities within the bed and out of bed (r = 0.622, p< 0.001). All items in the affective subscale were moderately correlated with each other between rho 0.421 – 0.483.

**Table Four:** Subscale item to total correlations and Cronbach alpha for a five-factor solution

|  | Scale<br>Mean if<br>Item<br>Deleted | Scale<br>Variance if<br>Item<br>Deleted | Corrected<br>Item-Total<br>Correlation | Cronbach's<br>Alpha if<br>Item<br>Deleted |
| --- | --- | --- | --- | --- |
| **Pain Severity and Interference Subscale (Cronbach alpha = 0.74)** |  |  |  |  |
| 1. On this scale please indicate the least pain that you had in the emergency department? | 27.62 | 81.776 | .661 | .639 |
| 2. On this scale please indicate the worst pain you had in the emergency department? | 24.69 | 111.506 | .363 | .748 |
| 3. How often were you in severe pain in the emergency department? | 26.28 | 83.370 | .439 | .726 |
| 4. Select one number below that best describes how much pain interfered or prevented you from: |  |  |  |  |
| A. Doing activities in bed such as turning, sitting up, repositioning | 25.92 | 81.306 | .513 | .693 |
| B. Doing activities out of bed such as walking, sitting in a chair, standing at sink | 26.05 | 72.834 | .620 | .647 |
| **Sleep subscale (Cronbach alpha = 0.86)** |  |  |  |  |
| 4. Select one number below that best describes how much pain interfered or prevented you from: |  |  |  |  |
| C. Falling asleep | 6.81 | 16.157 | .757 | . |
| D. Staying asleep | 6.84 | 15.501 | .757 | . |
| **Affective subscale (Cronbach alpha = 0.71)** |  |  |  |  |
| 5. Pain can affect our mood and emotions. On this scale, please select the one number that best shows how much the pain caused you to feel: |  |  |  |  |
| A. Depressed | 6.64 | 42.900 | .514 | .648 |
| B. Frightened | 6.97 | 43.245 | .523 | .638 |
| C. Helpless | 5.56 | 38.555 | .564 | .586 |
| **Side-effects subscale (Cronbach alpha = 0.55)** |  |  |  |  |
| 6. Have you had any of the following side effects? Please select "0" if no; if yes, please circle the number that best shows the severity of each. |  |  |  |  |
| A. Nausea | 4.32 | 34.694 | .312 | .502 |
| B. Drowsiness | 4.18 | 34.110 | .344 | .468 |
| C. Itching | 6.18 | 50.199 | .290 | .526 |
| D. Dizziness | 5.00 | 36.329 | .441 | .380 |
| **Perceptions of Care subscale (Cronbach alpha = 0.50)** |  |  |  |  |
| 7. In the emergency department, how much relief of your pain did you receive? | 1.87 | 12.017 | .280 | .516 |
| 8. Were you allowed to participate in decisions about your pain treatment as much as you wanted to? | 5.36 | 17.318 | .247 | .501 |
| 9. Select one number that best shows how satisfied you are with the results of your pain treatment in the emergency department. | 5.02 | 15.671 | .477 | .194 |

### Additional Construct Validity Testing

#### Contrasting Groups

Table Five shows the mean scores for satisfaction and the participant’s reports of their involvement in decisions regarding pain care in the different contrasting groups.

**Table Five:** Contrasting groups and patient satisfaction and involvement in their own pain care.

| Contrasting Group |  | Mean (95% CI)<br>Satisfaction<br>Score+ | Significance | Mean (95% CI) Participation<br>in decision about pain<br>treatment+ | Significance |
| --- | --- | --- | --- | --- | --- |
| Sex | Male | 0.96 (0.58 – 1.34) | 0.393 | 0.92 (0.38 – 1.46) | 0.835 |
|  | Female | 1.19 (0.80 – 1.59) |  | 0.65 (0.27 – 1.02) |  |
| Age | Below 65 years | 1.03 (0.75 – 1.32) | 0.899 | 0.71 (0.37 – 1.04) | 0.354 |
|  | Above 65 years | 1.04 (0.17 – 1.90) |  | 1.19 (0.14 – 2.25) |  |
| ISRAD | Disadvantaged | 1.61 (0.68 – 2.53) | 0.187 | 1.24 (0.21 – 2.28) | 0.287 |
|  | Advantaged | 0.92 (0.65 – 1.19) |  | 0.68 (0.35 – 1.00) |  |
| Private Health Insurance | Yes | 1.14 (0.71 – 1.57) | 0.763 | 0.80 (0.30 – 1.31) | 0.895 |
|  | No | 1.05 (0.69 – 1.41) |  | 0.75 (0.33 – 1.16) |  |
| Charlson Comorbidity Index | No Comorbidities | 1.10 (0.71 – 1.50) | 0.498 | 0.67 (0.26 – 1.08) | 0.503 |
|  | Comorbidities Present | 0.96 (0.58 – 1.33) |  | 0.88 (0.38 – 1.38) |  |
| Mode of Arrival | Walk-in | 1.04 (0.67 – 1.42) | 0.720 | 0.97 (0.48 – 1.46) | 0.133 |
|  | Ambulance Service | 1.14 (0.74 – 1.54) |  | 0.52 (0.16 – 0.97) |  |
| Australasian Triage Score | Urgent | 0.54 (0.20 – 0.89) | 0.015* | 0.59 (0.00 – 1.23) | 0.551 |
|  | Less Urgent | 1.11 (0.80 – 1.42) |  | 0.80 (0.44 – 1.16) |  |
| Analgesia Administered | Yes | 0.98 (0.71 – 1.25) | 0.515 | 0.75 (0.43 – 1.07) | 0.744 |
|  | No | 1.45 (0.20 – 2.70) |  | 0.95 (0.00 – 2.32) |  |
| Analgesia Within 30 min | Yes | 0.86 (0.29 – 1.43) | 0.545 | 0.61 (0.04 – 1.18) | 0.539 |
|  | No | 1.07 (0.76 – 1.38) |  | 0.81 (0.44 – 1.18) |  |

+ Lower scores indicate higher satisfaction with care and greater involvement in decision making surrounding the participants care.

There was a statistically significant difference in satisfaction scores between patients with an urgent triage score (ATS category 1 or 2) and those with a less urgent score (ATS category 3, 4 or 5). There were no other differences between contrasting groups.

## 7. Discussion

This study shows that it is feasible to use a modified version of the APS-POQ-R to assess patient-reported outcomes of pain care in the ED. The APS-POQ-R, modified for ED use, has demonstrated construct validity with five subscales. The APS-POQ-R has been validated in several different populations including general medical/surgical patients^20,22,25^ general surgical patients^23,38^ and patients with acute abdominal pain^24,39^, but this is the first time the English version has had construct validation demonstrated in the adult ED. However, further multi-centre testing is required before the widespread use of the instrument can be recommended.

Adult ED patients were generally agreeable to completing the modified APS-POQ-R upon completion of their ED care, with a recruitment rate of over 75%. As expected, the item-response rate was high with 92% of participants completing the instrument in full. Despite this, we expect that further content validation and refining to make the questions even more relevant to the ED will increase the item-response rate.

Construct validity of the instrument was demonstrated in the ED with all 17 items mapping to five subscales: Pain Severity and Interference with Activity subscale, Sleep subscale, Affective subscale, Side-effects subscale, and Perceptions of Care subscale. There are subtle differences between these subscales and those previously reported^20^. Gordon et al. found that items relating to pain intensity grouped together with items relating to sleep, and items relating to interference with activity sat in a separate subscale^20^. In contrast, in the current study, items about pain intensity group together with interference with activity items, and items relating to sleep mapped to their own subscale. The reasons for these differences are unknown but may reflect between-study differences in types of patients and the characteristics of their pain. It is reasonable to assume that the circumstances surrounding acute pain that makes a person present to ED are likely to differ from the circumstances surrounding pain experienced by hospitalized patients on medical and surgical wards. We can only speculate that ED patients tend to associate pain intensity with limiting of their usual activities, but that hospital inpatients associate pain intensity with limitations on sleep. The reasons leading to patients presenting to the ED in pain need further investigation and may inform the future revision of the pain severity and interference subscale.

Previous work has reviewed the patient-reported outcomes used in ED pain research^12^. In this scoping review, five areas of patient-reported outcome measurement were identified within the 56 studies included. These five areas of patient-reported outcomes map to the five subscales identified in the initial validation of the APS-POQ-R and now also map to the five constructs identified in the ED validation. The table below (Table Six) identifies these patient-reported outcomes.

**Table Six:** Comparisons of the patient-reported outcomes of care identified by Wong et al. 2020, the original APS-POQ-R and the ED validation of the APS-POQ-R

|  | Scoping Review | APS-POQ-R | Modified APS-POQ-R |
| --- | --- | --- | --- |
| 1. | Pain Intensity | Pain Severity and Sleep Interference | Pain Severity and Activity Interference |
| 2. | Patient Satisfaction | Perceptions of Care | Perceptions of Care |
| 3. | Side Effects | Adverse Effects | Side Effects |
| 4. | Emotional Functioning | Affective Functioning | Affective Functioning |
| 5. | Physical Functioning | Activity Interference | Sleep Interference |

The urgency of the presenting problem was discriminatory for reported patient satisfaction. Patients with a higher urgency (ATS 1 or 2) provided a more positive satisfaction score than patients in lower urgency categories (ATS 3 to 5). In previous work, the time-to-be-seen by a treating clinician has had a significant impact on metrics of pain care in the ED^14,33^ and urgency is a surrogate measure of time-to-be-seen by a treating clinician in the ED. The influence of the ED environment was not captured in this work therefore some of the discriminatory abiliy of this instrument may be missed. Further work into the use of the instrument and the determinants of patient-reported outcomes should take into account the impact of workload as this is a significant predictor of care in the ED environment, and has previously been shown to influence the treatment of pain within a symptom management model^18^.

## 8. Limitations

There are several limitations to this work. While the concepts included in this instrument are robust to pain care in other settings, it is possible that there are unique challenges related to pain care in the ED that is not covered by this instrument. The transcription error leading to an item being missed will have to be accounted for in future testing of the instrument, however, as the other affective questions mapped as expected, then we would also expect the omitted question about anxiety to map similarly. This study was a convenience sample of patients, recruited during business hours, Monday to Friday, who were able to consent and answer the questions in English. This means that patients in some groups (vulnerable, cognitively impaired, non-English-speaking) who are traditionally thought to receive poor ED pain care were excluded from the study. This may affect the applicability of the instrument to the broader ED population, however the current study represents a starting point towards a more comprehensive tool. This study was only undertaken in one ED, and before widespread use, confirmatory factor analysis in multiple EDs should occur.

## 9. Conclusion

The modified APS-POQ-R demonstrates construct validity for use in acute pain in the adult ED. This instrument covers all areas of the patient-reported outcomes of pain care currently described in ED pain care research in one instrument. This instrument is feasible to use in research and quality improvement in the ED environment. However, before the widespread use of the instrument, it should be further validated in a variety of ED settings.

## Data Availability

Data from this study is not available as the patients recruited did not give permission for data sharing.

